# Factors associated with participation over time in the Avon Longitudinal Study of Parents and Children: a study using linked education and primary care data

**DOI:** 10.1101/2020.03.10.20033621

**Authors:** RP Cornish, J Macleod, A Boyd, K Tilling

## Abstract

**Background:** Dropout from studies can lead to biased exposure-outcome estimates if the outcome is associated with continued participation, but this cannot be investigated using incomplete data. Linkage to external datasets provides a means of obtaining outcome – or proxy outcome - data on non-responders.

**Methods:** We examined the association between baseline socio-demographic factors and participation in the Avon Longitudinal Study of Parents and Children. We investigated whether child and adolescent outcomes measured in linked education and primary care data were associated with participation after accounting for baseline factors. To demonstrate the potential for bias, we examined whether the association between maternal smoking and these outcomes differed in the subsample who completed the 19-year questionnaire.

**Results:** Lower levels of school attainment, lower GP consultation and prescription rates, higher BMI, special educational needs (SEN) status, not having an asthma diagnosis, depression and being a smoker were all associated with lower participation after adjustment for baseline factors. For example, adjusted odds ratio (OR) for participation comparing ever smokers (by 18 years) to non-smokers: 0.65, 95% CI (0.56, 0.75). The association of maternal smoking differed between the subsample of participants at 19 years and the entire sample, although differences were small and confidence intervals overlapped. For example: for SEN status OR=1.19 (1.06, 1.33) (all participants); OR=1.03 (0.79, 1.45) (subsample).

**Conclusions:** Linkage to routine data provides a unique opportunity to compare continuing participators to those who drop out, and the impact this self-selection can have on results. Cohort studies should use linkage to routine data to explore participation and conduct sensitivity analyses.

**Key messages:**

- Educational and health-related characteristics are strongly associated with ongoing participation in the Avon Longitudinal Study of Parents and Children after adjustment for detailed socio-demographic factors.
- This could bias analyses using the dataset, with bias dependent on the variables used in the analysis and their impact on participation.
- Linkage to routine data provides a means of assessing whether factors measured across the life course are associated with ongoing participation in observational studies and the potential impact of this in terms of bias.
- Researchers can use linkage to external sources of data to make informed decisions about the likely impact of selective participation and to guide their analyses.

## Introduction

Dropout from longitudinal studies (we will refer to not dropping out as participation) will bias estimates of exposure-outcome associations if the association between the exposure and the probability of participation is differential with respect to the outcome, or vice versa [1]. The circumstances under which this occurs depends on the analysis model being used [2]. To know whether a given analysis model is likely to be biased by dropout, we need to be able to assess whether exposure and outcome are related to participation – this is impossible to do using only the observed study data. However, complete data on some post-baseline variables may be obtained via linkage to external datasets. As with any observational study, associations between a factor and participation may be causal (smoking may cause dropout), or confounded (lower education levels may cause both dropout and smoking). One way to address this is by using genetic information [3, 4]. A recent study using Avon Longitudinal Study of Parents and Children (ALSPAC) data, [3] found that, in both the mothers and index children, polygenic scores for years of education and agreeableness were associated with greater participation; conversely, polygenic scores for smoking initiation, schizophrenia, ADHD and depression were associated with lower levels of participation.However, using genetic information has several disadvantages. Firstly, the genetic contribution to phenotypes is often low, limiting power; secondly, not all factors of interest can be genetically instrumented; and, finally, genetic information may not be available for everyone.

Here we aimed to complement previous studies examining genetic predictors of participation by examining associations with observed phenotypes as recorded in linked education or primary care data as well as with factors that cannot (or have not) been genetically instrumented (school absence, special educational needs classification, General Practitioner (GP) consultation rates, counts of prescribed drugs). We examine factors associated with continuing participation in ALSPAC, but the methods are applicable to any cohort study or long-term follow up of a clinical trial.

## Methods

### Sample/subjects

ALSPAC is a prospective study – described in detail previously [5, 6] – which recruited pregnant women living in and around Bristol, south west England, with due dates between April 1991 and December 1992. Initially, 14 541 pregnant women enrolled, resulting in 14 062 live births and 13 988 infants alive at one year. Detailed data were collected during pregnancy and participants have been followed up since birth through questionnaires, clinics and linkage to routine datasets. Ethical approval was obtained from the ALSPAC Ethics and Law Committee and Local Research Ethics Committees: http://www.bristol.ac.uk/alspac/researchers/research-ethics/. (Note that ALSPAC has a searchable data dictionary and variable search tool: http://www.bristol.ac.uk/alspac/researchers/our-data/).

We used data from singletons and twins who were alive at one year and had not subsequently withdrawn (n=13 972).

### ALSPAC participation

We defined participation as returning a completed questionnaire or attending a study assessment clinic. Participants in this study were mothers or carers (henceforth mothers), who completed questionnaires about themselves or their child, and the (index) children, who (from age 65 months) also completed questionnaires. Clinics before age 7 years were aimed at a subset of enrolled children; we only considered questionnaires and clinics (up to age 20) that all participants were eligible to complete/attend. We have considered mother-completed questionnaires (about themselves or their child) separately from clinics and child-completed questionnaires; the latter two constitute *child participation*, the former *mother participation*. A complete list of questionnaires and clinics that we included is given in Supplementary Table 1.

### Baseline socio-demographic variables

Baseline socio-demographic and other variables potentially associated with participation were included. The majority of these were factors measured in pregnancy, since this was when response rates were highest. The variables were: maternal ethnicity, age, parity, marital status, age at first pregnancy, educational level, smoking, and depression score (Edinburgh Postnatal Depression Scale at 18 weeks gestation); housing tenure; whether the mother or her partner had use of a car; whether their house had double glazing; whether they had a phone in their home; number of rooms in the house; crowding index, defined as number of people per room (excluding bathrooms and toilets); family occupational social class, defined as the higher of maternal and paternal social class and categorised as I-IIIN (professional, managerial, and non-manual skilled occupations) and IIIM-IV (manual skilled, semi-skilled and unskilled occupations); duration of breastfeeding, derived from responses to questionnaires administered at 4 weeks, 6 months and 15 months; and child sex. Paternal factors were not included – because response rates for these were lower and because, in a preliminary analysis, none of the paternal factors considered (education, smoking, and depression score) were associated with participation after taking account of maternal and other factors listed above.

### Education variables from the National Pupil Database (NPD)

The NPD is a database containing attainment and other data for children attending schools in England (https://www.gov.uk/government/collections/national-pupil-database). Linkage between ALSPAC and the NPD has been described previously [7]. Here we used three variables from Year 11 (age 15-16 years): capped point score (a measure of attainment), described previously [7]; percent attendance; and special educational needs (SEN) status, classified as none, school action support, or SEN Statement. [SEN Statements – now replaced by Education, Health and Care Plans (EHC Plans) –are a description of a child’s educational needs and any additional support they should receive in school.]

### Linkage to primary care data

When the index children reached legal adulthood (age 18), ALSPAC conducted a postal fair processing campaign to re-enrol them into the study in their own right and to seek (opt-out) permission for linkage to health and administrative records. Linkage to primary care records was carried out following this campaign and is described in the supplementary material.

### Variables derived from linked primary care data

Adolescent BMI: the mean of all recorded measurements after age 10.

Consultation rate age 15-19 years: the total number of consultations during this period, divided by five.

Mean (prescribed) drug count age 15-19 years: total drug count in this period divided by five. Asthma diagnosis: Read code for a diagnosis [8] before 8 years.

Depression: Read code for a diagnosis, symptoms or treatment before the age of 18 ([9]). Smoking: based on Read codes used in two recent studies [10, 11]; we classified individuals as having a record for ever or current smoking (or not) before age 18 (supplementary material).

### Numbers with linked data

Of the 13 972 individuals included in this analysis, 12 395 (89%) were sent fair processing materials. Of these, 360 (3%) dissented to linkage to education records and 423 (3%) to health records. Of the remaining 12 035, 11 414 were linked to the NPD and had data on at least one of the variables used in this analysis. Of the 11 972 who did not dissent to health data linkage, ALSPAC had no NHS ID for 17, leaving 11 955 where linkage to primary care records was attempted. Primary care records (not necessarily for the entire time period) were extracted for 11 087 (93% of individuals where linkage was possible; 79% of the original 13 972).

### Statistical analysis

For each questionnaire and study clinic, a binary variable was created to indicate whether each individual participated (returned that questionnaire or attended the clinic). Two random effects logistic regression models (one for mother and one for child participation) were used to model participation over time using cubic splines [12], with 5 knots placed using Stata’s default method [13]. We used the (fixed) age at which the questionnaire/clinic invitation was sent, rather than the actual age at completion/attendance. For the mother-completed questionnaires, we used time in study, where time in study = 0 denotes the beginning of pregnancy.

Multiple imputation using chained equations [14] was used to impute missing data. Two models were used. The first imputation model (which included all 13 972 individuals) was used to impute baseline covariates and linked education variables; the model included the baseline covariates, the three linked education variables (attainment score, percent absence, and SEN status), and all binary participation variables (both mother and child). IQ measured when the children were aged 8 using the Wechsler Intelligence Scale for Children (WISC-III) [15] was included as an auxiliary variable. A second imputation model was used to examine the association between measures based on primary care data and participation. This model only included individuals with primary care data beyond the age of four (n=10 811) and used baseline covariates, all participation variables, the seven GP measures and, as auxiliary variables, consultation rates and drug counts at additional ages (0-4 years, 5-9 years, 10-14 years and 20+ years). Further details regarding missing data and the imputation models are given in the supplementary material. As a sensitivity analysis, we carried out a complete case analysis.

To investigate the impact of selective participation on exposure-outcome estimates we used linear and logistic regression (as appropriate) to examine the association between maternal smoking (ever vs never, measured in early pregnancy) and the following outcomes: attainment score, percent school absence, SEN status (dichotomised: school action support/statement of SEN vs none), asthma, BMI, depression, and smoking. The numerical variables – attainment, school absence and BMI were converted to z-scores (SD units) for this analysis. These associations were examined (in the imputed data) among (1) all individuals and (2) among individuals who completed the ALSPAC questionnaire administered at 19 years (the latest child-completed questionnaire included in our analysis). As a sensitivity analysis, this was repeated among the complete cases (i.e. all those with data on maternal smoking, the outcome of interest, plus baseline covariates) then among the subset of these who completed the age 19 questionnaire.

Analyses were carried out in Stata 15 [16].

## Results

Participation rates by the mother were high in pregnancy and gradually declined over time, particularly when the children reached mid-late adolescence (Figure 1). Child participation remained stable during childhood (children started completing questionnaires at around 5 years of age) but declined during adolescence.

**Figure 1:**
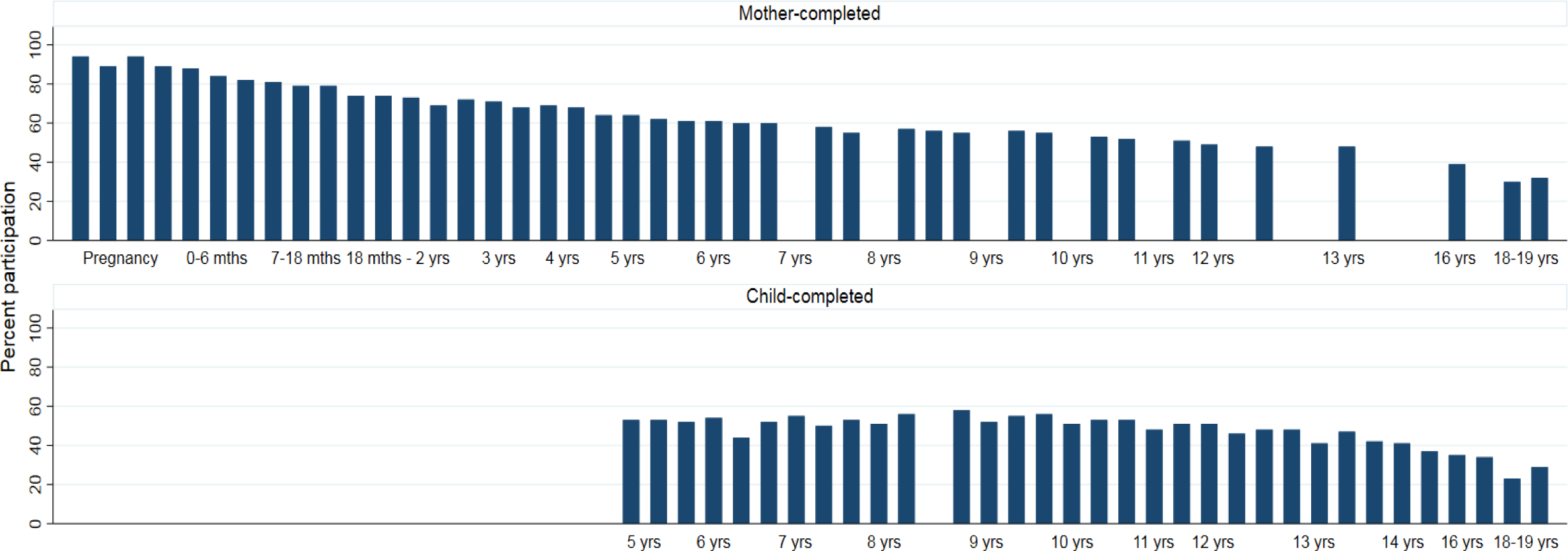
Participation rates (%) in ALSPAC: mother and child completed Enrolment in ALSPAC is defined as the participant having actively participated at least once. However, no single assessment has complete coverage as there was no single baseline assessment administered to all participants, given that mothers were recruited at different stages of pregnancy or following delivery, and may therefore have missed an earlier assessment.

Baseline covariate data were missing for between 0% (sex and mother’s age at birth) and 18% (family occupational social class) of the 13 972 individuals (Supplementary Table S1) and 9049 (65%) had complete covariate information. Individuals with complete baseline covariates had higher levels of participation than the overall sample (mean number of mother-completed questionnaires was 31, compared to 27 in the overall sample; mean number of child-completed questionnaires/clinics attended was 19, compared to 16 in the overall sample).

### Predictors of participation: baseline variables

Table 1 gives mutually adjusted odds ratios (ORs) for child and mother participation (obtained using multiple imputation (MI)) for all baseline covariates. ORs for child and mother participation were very similar, with the exception of sex (of child), which was a strong predictor of child participation but not of mother participation. The ORs from the complete case analysis were generally similar to those obtained using MI (Supplementary Table S2), although ORs for breastfeeding and maternal ethnicity were less extreme in the complete case analysis.

**Table 1:** Odds ratios for participation for all baseline covariates (50 imputed datasets; n=13 972)

| Covariate | Level | Child participation OR (95% CI) | p-value | Mother participation OR (95% CI) | p-value |
| --- | --- | --- | --- | --- | --- |
| Child sex | Female vs male | 1.88 (1.71, 2.06) | <0.001 | 1.10 (0.99, 1.22) | 0.07 |
| Mother's education | O level / lower | 1.00 |  | 1.00 |  |
|  | A level | 1.47 (1.28, 1.68) |  | 1.58 (1.35, 1.85) |  |
|  | Degree/higher | 1.66 (1.38, 2.00) | <0.001 | 1.74 (1.40, 2.18) | <0.001 |
| Parity | 0 | 1.00 |  | 1.00 |  |
|  | 1 | 0.81 (0.70, 0.93) |  | 0.84 (0.72, 0.99) |  |
|  | 2+ | 0.59 (0.48, 0.73) | <0.001 | 0.60 (0.48, 0.76) | <0.001 |
| Mother's age (at birth of index child) | Per 1 year increase | 1.07 (1.06, 1.09) | <0.001 | 1.07 (1.05, 1.08) | <0.001 |
| Mother's ethnicity | Non-white vs white | 0.26 (0.19, 0.36) | <0.001 | 0.14 (0.10, 0.19) | <0.001 |
| Family social class | Manual vs non-manual | 0.73 (0.63, 0.85) | <0.001 | 0.63 (0.52, 0.75) | <0.001 |
| Age at first pregnancy | <20 | 1.00 |  | 1.00 |  |
|  | 20-24 | 1.36 (1.17, 1.59) |  | 1.32 (1.11, 1.56) |  |
|  | 25+ | 1.63 (1.37, 1.95) | <0.001 | 1.85 (1.51, 2.25) | <0.001 |
| Maternal smoking | Yes vs no (in pregnancy) | 0.80 (0.69, 0.93) | 0.003 | 0.80 (0.67, 0.94) | 0.008 |
|  | Yes vs no (ever) | 0.78 (0.68, 0.88) | <0.001 | 0.81 (0.71, 0.93) | 0.003 |
| Duration of breastfeeding | Never/<1 month | 1.00 |  | 1.00 |  |
|  | 1 to <3 months | 2.99 (2.56, 3.51) |  | 3.74 (3.12, 4.48) |  |
|  | 3 to <6 months | 2.69 (2.26, 3.20) |  | 2.92 (2.37, 3.61) |  |
|  | 6 months+ | 3.97 (3.45, 4.56) | <0.001 | 5.27 (4.46, 6.23) | <0.001 |
| Married | Yes vs no | 1.33 (1.16, 1.53) | <0.001 | 1.39 (1.19, 1.62) | <0.001 |
| Housing tenure | Owned/mortgaged | 1.00 |  | 1.00 |  |
|  | Private rented | 0.50 (0.40, 0.63) |  | 0.49 (0.39, 0.63) |  |
|  | Council/HA/other | 0.69 (0.59, 0.82) | <0.001 | 0.66 (0.55, 0.79) | <0.001 |
| Number of rooms | Per 1 room increase | 1.02 (0.97, 1.07) | 0.5 | 1.00 (0.94, 1.06) | 1.0 |
| Phone in home | Yes vs no/incoming only | 0.66 (0.54, 0.79) | <0.001 | 0.68 (0.55, 0.84) | <0.001 |
| Car use | No vs yes | 0.53 (0.44, 0.65) | <0.001 | 0.61 (0.49, 0.76) | <0.001 |
| Double glazing | None vs full/partial | 0.88 (0.79, 0.98) | 0.02 | 0.86 (0.76, 0.96) | 0.009 |
| Financial difficulties | Per 1 unit increase | 0.98 (0.96, 0.99) | 0.008 | 0.97 (0.95, 0.99) | 0.004 |
| Crowding index | ≤0.5 | 1.00 |  | 1.00 |  |
|  | >0.5 – 0.75 | 0.93 (0.80, 1.08) |  | 0.85 (0.72, 1.01) |  |
|  | >0.75 – 1 | 0.85 (0.70, 1.03) |  | 0.79 (0.62, 0.99) |  |
|  | >1 | 0.75 (0.57, 1.00) | 0.2 | 0.59 (0.43, 0.81) | 0.06 |
| Depression score | Per 1 unit increase | 0.98 (0.97, 0.99) | 0.004 | 0.98 (0.97, 0.99) | 0.001 |

### Predictors of participation: child education variables

After adjusting for baseline factors, all three education variables were associated with child participation (Table 2). Those with higher attainment scores, lower absence rates and no recorded SEN were more likely to participate. Attainment and absence (but not SEN status) were also associated with mother participation. Results were similar in the complete case analysis (Supplementary Table S3).

**Table 2:**
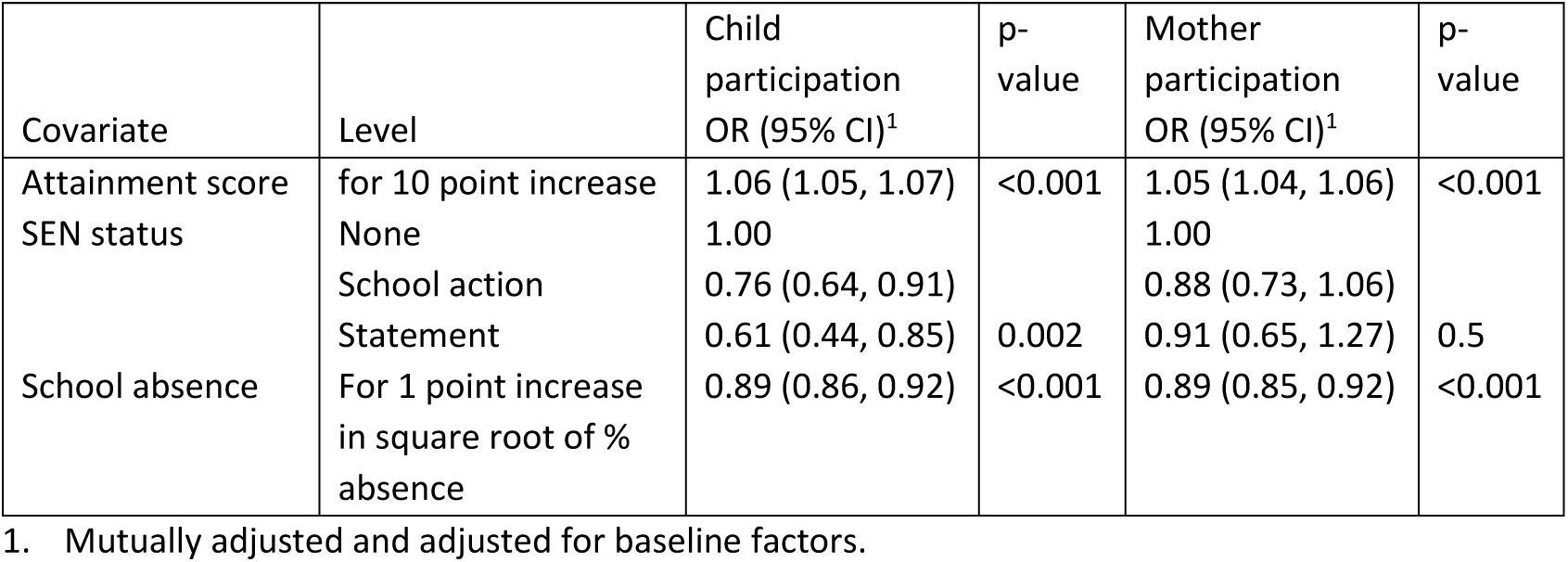
Odds ratios for participation for education variables (50 imputed datasets; n=13 972)

| Covariate | Level | Child participation OR (95% CI)<sup>1</sup> | p-value | Mother participation OR (95% CI)<sup>1</sup> | p-value |
| --- | --- | --- | --- | --- | --- |
| Attainment score | for 10 point increase | 1.06 (1.05, 1.07) | <0.001 | 1.05 (1.04, 1.06) | <0.001 |
| SEN status | None | 1.00 |  | 1.00 |  |
|  | School action | 0.76 (0.64, 0.91) |  | 0.88 (0.73, 1.06) |  |
|  | Statement | 0.61 (0.44, 0.85) | 0.002 | 0.91 (0.65, 1.27) | 0.5 |
| School absence | For 1 point increase in square root of % absence | 0.89 (0.86, 0.92) | <0.001 | 0.89 (0.85, 0.92) | <0.001 |

1. Mutually adjusted and adjusted for baseline factors.

### Predictors of participation: child measures derived from primary care data

After adjusting for baseline factors, all primary care measures were related to child participation, although the association with BMI was weak (Table 3). GP-recorded (child) asthma diagnosis by age 8 and smoking before age 18 were both strongly associated with participation by the mother. The ORs for the association between the other GP-derived variables and mother participation were in the same direction (but smaller than) those for child participation, with confidence intervals including the null. The complete case associations were generally larger in magnitude but in the same direction as those obtained using MI; the exception to this was for asthma (Supplementary Table S5).

**Table 3:**
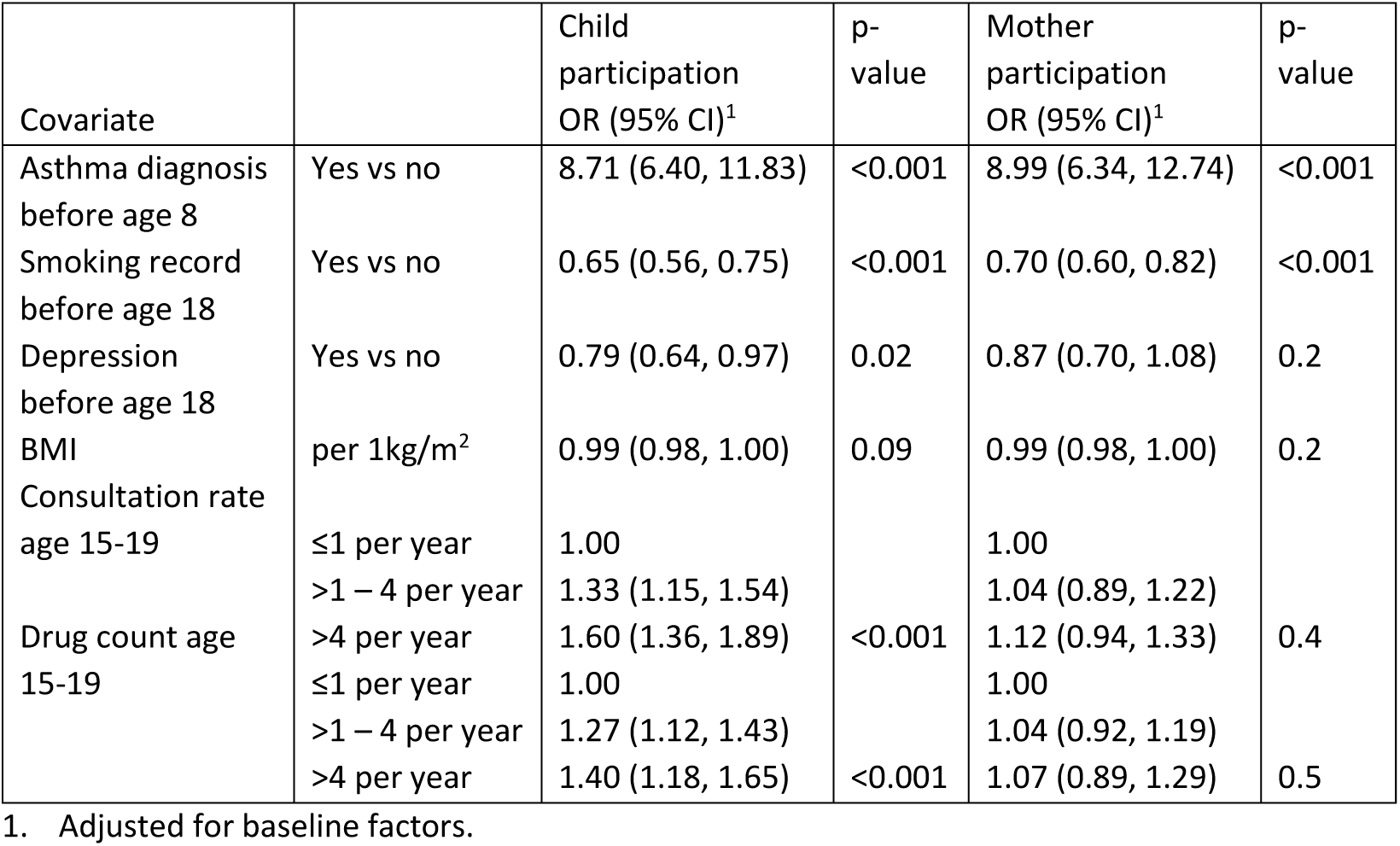
Odds ratios for participation for GP-derived child measures (50 imputed datasets: n=10 811)

| Covariate |  | Child participation OR (95% CI)<sup>1</sup> | p-value | Mother participation OR (95% CI)<sup>1</sup> | p-value |
| --- | --- | --- | --- | --- | --- |
| Asthma diagnosis before age 8 | Yes vs no | 8.71 (6.40, 11.83) | <0.001 | 8.99 (6.34, 12.74) | <0.001 |
| Smoking record before age 18 | Yes vs no | 0.65 (0.56, 0.75) | <0.001 | 0.70 (0.60, 0.82) | <0.001 |
| Depression before age 18 | Yes vs no | 0.79 (0.64, 0.97) | 0.02 | 0.87 (0.70, 1.08) | 0.2 |
| BMI | per 1kg/m<sup>2</sup> | 0.99 (0.98, 1.00) | 0.09 | 0.99 (0.98, 1.00) | 0.2 |
| Consultation rate age 15-19 | ≤1 per year | 1.00 |  | 1.00 |  |
|  | >1 – 4 per year | 1.33 (1.15, 1.54) |  | 1.04 (0.89, 1.22) |  |
| Drug count age 15-19 | >4 per year | 1.60 (1.36, 1.89) | <0.001 | 1.12 (0.94, 1.33) | 0.4 |
|  | ≤1 per year | 1.00 |  | 1.00 |  |
|  | >1 – 4 per year | 1.27 (1.12, 1.43) |  | 1.04 (0.92, 1.19) |  |
|  | >4 per year | 1.40 (1.18, 1.65) | <0.001 | 1.07 (0.89, 1.29) | 0.5 |

1. Adjusted for baseline factors.

### Impact of non-response on exposure-outcome associations

Figure 2 gives estimated associations (ORs or regression coefficients) between maternal smoking (ever/never) and outcomes derived from linked data for all individuals and for the subset who completed the questionnaire at 19 years. The associations in the subsample were all in the same direction as in the full sample and some were of a similar magnitude. The associations with offspring depression and smoking were stronger in the subsample; conversely, the associations with SEN status, asthma and attainment were attenuated in the subsample, although the confidence intervals overlapped in every case.

**Figure 2:**
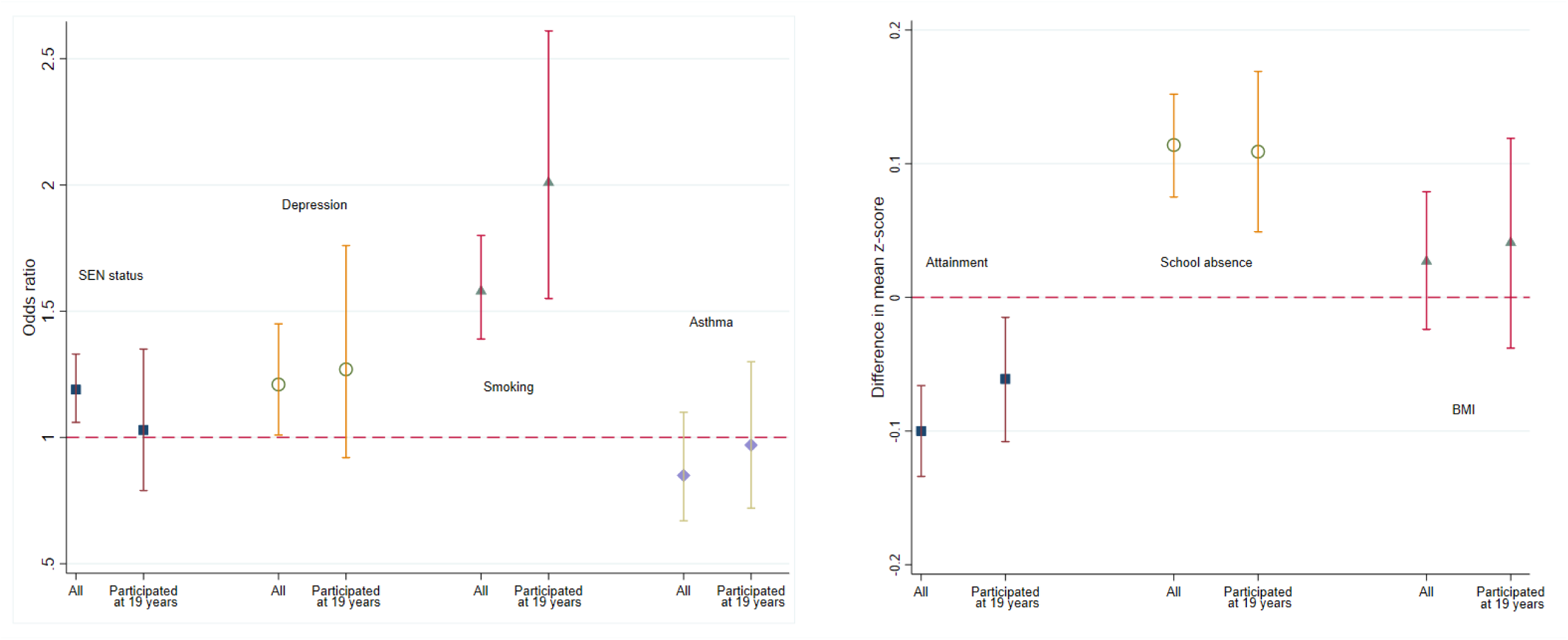
Association between maternal smoking (y vs no) and various adolescent outcomes

## Discussion

We found that participation in ALSPAC is associated with a wide range of baseline socio-demographic factors; in all cases, factors suggesting greater social disadvantage were associated with lower participation. After taking account of these factors, higher educational attainment, asthma diagnosis and higher GP consultation rates and prescribed drug counts were associated with higher participation; greater school absence, special education needs status, smoking, depression, and higher BMI were associated with lower participation. We also showed that some exposure-outcome estimates may be biased if restricted to current participants.

Recently, others found evidence that polygenic risk scores for education, smoking initiation, BMI, and depression – as well as scores for other traits – are associated with ongoing participation in ALSPAC [3]; these associations were in the same direction as those found in the current study. Similar findings have been reported in a review of factors associated with participation in epidemiologic studies [17] and in studies looking specifically at the association between psychiatric disorders and dropout in longitudinal studies [18, 19]. Although some have suggested that the associations with participation would have to be quite extreme for the resulting bias to be of concern [20], results from non-randomised studies are increasingly being combined and used to inform clinical practice [21, 22]. Even a relatively small amount of bias could be problematic, particularly if similar characteristics are related to ongoing participation in different studies – results may then be reproducible from one study to another, but these results may be equally biased. In 1998, Egger et al. discussed the pitfalls of combining results from observational studies. In particular, they argued that such studies may give rise to findings that are biased and combining these (biased) estimates will result in an overall finding that is “very precise but equally spurious” [23].

A key limitation of our study is the fact that most of the variables were incomplete. By definition, the group with complete data had higher participation rates than those with one or more missing covariates. This may mean that the observed associations with participation are themselves biased. Similarly, linked data were not available for all individuals and these individuals are not a random subsample of the cohort. Individuals who had died, withdrawn from the study after age 14, could not be traced, or were flagged on the ALSPAC administrative database as being not contactable were not sent fair processing materials and were thus not eligible to be included in the linkage. Most of those without linked education data are those who were attending an independent (fee paying) school at the time of linkage [7]. Likewise, since the majority of primary care data come from local practices, the sample with linked GP data will mainly comprise those who have not moved out of the area. In addition, among individuals with linked primary care data, those with either low or high BMI may be more likely to have it recorded (because there may be health concerns about adolescents who appear obviously over or underweight). This could result in bias in the estimate of the association between BMI and participation. We expect the estimates of associations with participation obtained using MI to be less biased than complete case analyses because, by including participation at each time point, all the factors from the linked data, as well as additional auxiliary variables from ALSPAC (IQ, for example) we have a better approximation to missing at random (MAR). However, the associations with participation obtained using MI may still be subject to bias.

Our findings have several implications. Statistical methods used to analyse incomplete data all make assumptions about the missing data mechanism which cannot be investigated using the incomplete data alone. For example, standard implementations of MI assume the data are MAR, conditional on the variables included in the imputation model. Similarly, a complete case analysis will generally produce unbiased estimates of the exposure-outcome association if missingness is unrelated to the outcome of interest [24]. We found that many baseline and later measures were associated with ongoing participation; this finding is unlikely to be unique to ALSPAC, as indicated recently [25]. This suggests that – firstly – a wide range of baseline covariates may need to be included in the (complete case) analysis – or imputation/weighting model – in order for the assumptions to be met. Secondly, outcomes measured in adolescence were associated with participation, suggesting that a complete case analysis for such outcomes is likely to be biased.

Our study illustrates two key advantages of linking to external datasets. Firstly, it is not usually possible to examine the impact of non-baseline factors on ongoing study participation because these factors are – by definition – not measured on individuals who have dropped out, whereas linkage to external datasets means later measures could be obtained on all (or a reasonably large proportion of) individuals in a study regardless of whether they remain active participants. Secondly, previous work suggests that the inclusion in MI models of auxiliary variables that are reasonably highly correlated with a missing outcome variable is likely to reduce bias [26], as it gives a closer approximation to MAR. As in the previous study, these auxiliary variables could be proxies for a missing study outcome.

In conclusion, we have shown using linked data that health and educational factors are associated with ongoing participation in ALSPAC. Use of linked data will help future researchers establish whether specific analyses are likely to be biased if restricted to complete cases, and which statistical methods are likely to minimise bias.

## Data Availability

The authors do not have the authority to share the data that support the findings of this study, due to ALSPAC data access permissions, but any researcher can apply to use ALSPAC data, including the variables used in this investigation. Information about access to ALSPAC data is given on their website: (http://www.bristol.ac.uk/alspac/researchers/access/).

## Funding

This work was supported by the Medical Research Council [MR/L012081] and the Wellcome Trust [WT086118/Z/08/Z]. The UK Medical Research Council (MRC), the Wellcome Trust and the University of Bristol currently provide core funding for ALSPAC [102215/2/13/2]. Data collection is funded from a wide range of sources. This publication is the work of the authors and Rosie Cornish will serve as guarantor for the contents of this paper. RC and KT work in the MRC Integrative Epidemiology Unit funded by the UK Medical Research Council (MC_UU_00011/3) and the University of Bristol.

## Authors’ contributions

RC and KT conceived the study. RC conducted the statistical analyses and wrote the first draft of the manuscript. KT contributed to the interpretation of the results. All authors contributed to the drafting and revising of the manuscript. All authors have read and approved the final version.

## Acknowledgements

We are extremely grateful to all the families who took part in this study, the midwives for their help in recruiting them, and the whole ALSPAC team, which includes interviewers, computer and laboratory technicians, clerical workers, research scientists, volunteers, managers, receptionists and nurses.

## Notes

### Competing Interest Statement

The authors have declared no competing interest.

